# An Evaluation of Progress Towards the 2035 WHO End TB Targets in 40 High Burden Countries

**DOI:** 10.1101/2020.10.02.20175307

**Authors:** Jaeyoon Cha, Guy E. Thwaites, Philip M. Ashton

## Abstract

Tuberculosis (TB) is the leading global cause of death from a single infectious agent, with more than 10 million new cases annually. As a part of its efforts to control TB, the World Health Organisation (WHO) adopted the End TB Strategy in 2014 to reduce TB incidence by 90% between 2015 and 2035, with intermediate targets every five years.

We examined TB incidence data from 2000 to 2018 for 40 high burden countries (HBCs) from the WHO published statistics, contextualized and assessed their trends over time, and projected the incidence to 2035 for each country. Two recurrent patterns accounted for 26 of the 40 HBCs: linear decrease (n = 14) or a peak in the 2000s followed by decline (n = 12). As uncontrolled HIV is the greatest risk factor for TB, trends in HIV infected and uninfected people were analysed separately for 15 Sub-Saharan African HBCs with high HIV prevalence.

The projections of current trends were compared against the reductions required to meet the WHO End TB targets. Of the 25 countries without a high burden of HIV, only 5 are on track to meet the End TB targets: Ethiopia, Laos, Myanmar, Russia, and South Korea. Of the 15 high HIV burden countries, 6 are on track: Eswatini, Kenya, Lesotho, South Africa, Tanzania, and Zimbabwe. Three high HIV burden countries, Botswana, Namibia, and Zambia will miss the End TB targets due to the diminishing returns of indirectly decreasing TB incidence by controlling the HIV epidemic.

Overall, we predict 62 million excess cases of TB between 2020 and 2035 in the 29 HBCs projected to miss the WHO End TB targets. In high HIV burden countries, new programs aimed directly at TB will be required to maintain momentum. Moreover, our projections are based on data prior to the COVID-19 pandemic; the disruption of the pandemic is overwhelmingly likely to interrupt vital TB services and increase TB incidence. We anticipate that these findings will help orientate countries to their progress towards the End TB goals and inform the level of investment required to meet these important targets for a TB-free world.

**Research in context:** *Evidence before this study:* We searched PubMed and Google Scholar with the terms “End TB assessment,” “End TB Strategy,” or “WHO End TB” between June 2019 and September 2020 to identify studies in English reporting on progress towards meeting the End TB targets. We only identified two studies that carried out a quantitative assessment of current estimates against the End TB targets. One study limited its analyses to current estimates, rather than using statistical methods to produce projections based on current trends, while the other study and the WHO Global TB Report 2019 presented projections to 2020 only. No report provided a per-country estimate of the progress toward the End TB targets through 2035.

*Added value of this study:* This study presents projections of TB incidence from 2020 to 2035 for 40 high TB burden countries. We benchmark the progress of each country against the reductions necessary to meet the WHO End TB targets. We provide per country (rather than per WHO region) breakdowns of these numbers and place the results into broader global health and socio-political contexts. Additionally, we separately model incidence trends in HIV infected and uninfected populations to account for different trajectories in the two populations.

*Implications of all the available evidence:* Only 11 of the 40 countries assessed are on track to meet the 2035 End TB targets, leading to a total of 62 million excess cases of TB compared with if the targets were met. This is consistent with previous reports such as the WHO Global TB Report 2019, which found that only 11 of 30 high burden countries were on track to meet targets for 2020. We additionally demonstrate that TB specific programs should be developed in most high HIV burden countries, as reductions in TB in HIV uninfected people are declining much more slowly than in HIV infected people.

## Introduction

*Mycobacterium tuberculosis*, the causative agent of tuberculosis (TB), claims more lives than any other infectious organism. In 2018 alone, there were 1.4 million deaths and 10.0 million new cases of TB worldwide, with the 30 highest burden countries (HBCs) bearing 87% of the incidence.^1^ Twenty-five of the 30 countries are classified as low or lower-middle income by the World Bank.

With HIV/AIDS, ^2,3^ smoking,^4^ diabetes mellitus,^5^ malnutrition,^5,6^ and air pollution^7^ among the risk factors, TB is a global health concern of many faces.^1^ Particularly, HIV-positive individuals are 20 to 30 times more likely to develop active TB infection.^2,3^ In Sub-Saharan Africa, where the prevalence of HIV/AIDS is the highest globally, up to 70% of TB patients are estimated to be coinfected with HIV.^3^ As coinfection is known to increase mortality, TB is an especially pressing epidemic in this region. The burden of TB further extends beyond mortality to severe productivity and treatment costs that exacerbate the difficult economic climates in many HBCs. In 2018, TB cost the global economy $1 billion US dollars in missed workdays from sickness, $11 billion from an average loss of 15-years’ income from TB deaths, and $9.2 billion for treatment.^8^

To support its vision of a world free of TB, the World Health Organisation (WHO) developed the End TB Strategy, which includes ambitious targets and milestones for 2020, 2025, 2030 and 2035, and a high-level strategy to achieve them. The targets include: 95% reduction in TB deaths between 2015 and 2035, 90% reduction in TB incidence in the same time span, and no families facing catastrophic costs as a result of TB.^9^ Accurate assessment of TB incidence is thus a prerequisite and critical part of the End TB Strategy. The WHO estimations of incidence for each country is tailored according to the information available, which depends on the strength of the country’s public health system and surveillance measures. While accurately assessing TB incidence in high burden countries without strong public health systems is challenging, it is reassuring that a study from the Global Burden of Disease consortium, using different methods than the WHO, found a similar overall TB incidence in 2015 at 10.2 million cases compared to the WHO’s 10.4 million.^10,11^

By the WHO’s own reckoning, “[of] the world as a whole, most WHO regions and many high TB burden countries are not on track to meet the 2020 milestones of the End TB Strategy”.^1^ Between 2015 and 2018, there was a cumulative reduction in global incidence by 6.3%, far short of the 20% milestone set by the End TB Strategy for 2020. ^1^ The WHO’s Global Tuberculosis Report 2019 estimates that only 11 of the 30 HBCs are on track to meet the 2020 milestone for incidence: Cambodia, Ethiopia, Kenya, Lesotho, Myanmar, Namibia, Russia, South Africa, Tanzania, Zambia, and Zimbabwe.^1^ Other studies have assessed this first milestone with grim results. A CDC (Centres for Disease Control and Prevention) report concluded that only the Europe region, which includes no countries on the HBC list, is likely to meet the 2020 incidence target.^12^ Another study by Pan et al. predicts that only 2 regions out of 21 (Sub-Saharan Africa and Eastern Europe) and a mere 11 countries of the 195 analysed are on track to meet the 20% decrease in incidence; of these, two countries, South Africa and Russia, are also evaluated in this present study.^13^

This study aims to characterise the current trends in TB incidence in HBCs and place them into the context of the incidence End TB milestones and targets through 2035. Our projections extend beyond the WHO’s analyses and previously published reports, which currently only project to 2020, and provide detailed country-level breakdowns and contextual analyses. We believe that this longer-term view, while subject to considerable uncertainty, will enable stakeholders of HBCs to more effectively advocate for the resources necessary to meet these targets.

## Methods

### Data gathering

Countries were selected for analysis based on the WHO list of HBCs for TB in the post-2015 era: 14 countries identified for TB, MDR-TB, and TB/HIV; 8 countries for TB and TB/HIV only; 6 countries for TB and MDR-TB only; 2 countries for overall TB only; and 8 countries for TB/HIV only (full list available in Table 1).^1^ In addition to these 38 countries from the WHO list, Laos and South Korea were included. Laos is an immediate neighbour of Vietnam where we are based, and South Korea is a notable example of a high income country with a high burden of TB that may serve as a model for rapidly developing countries.

**Table 1.**
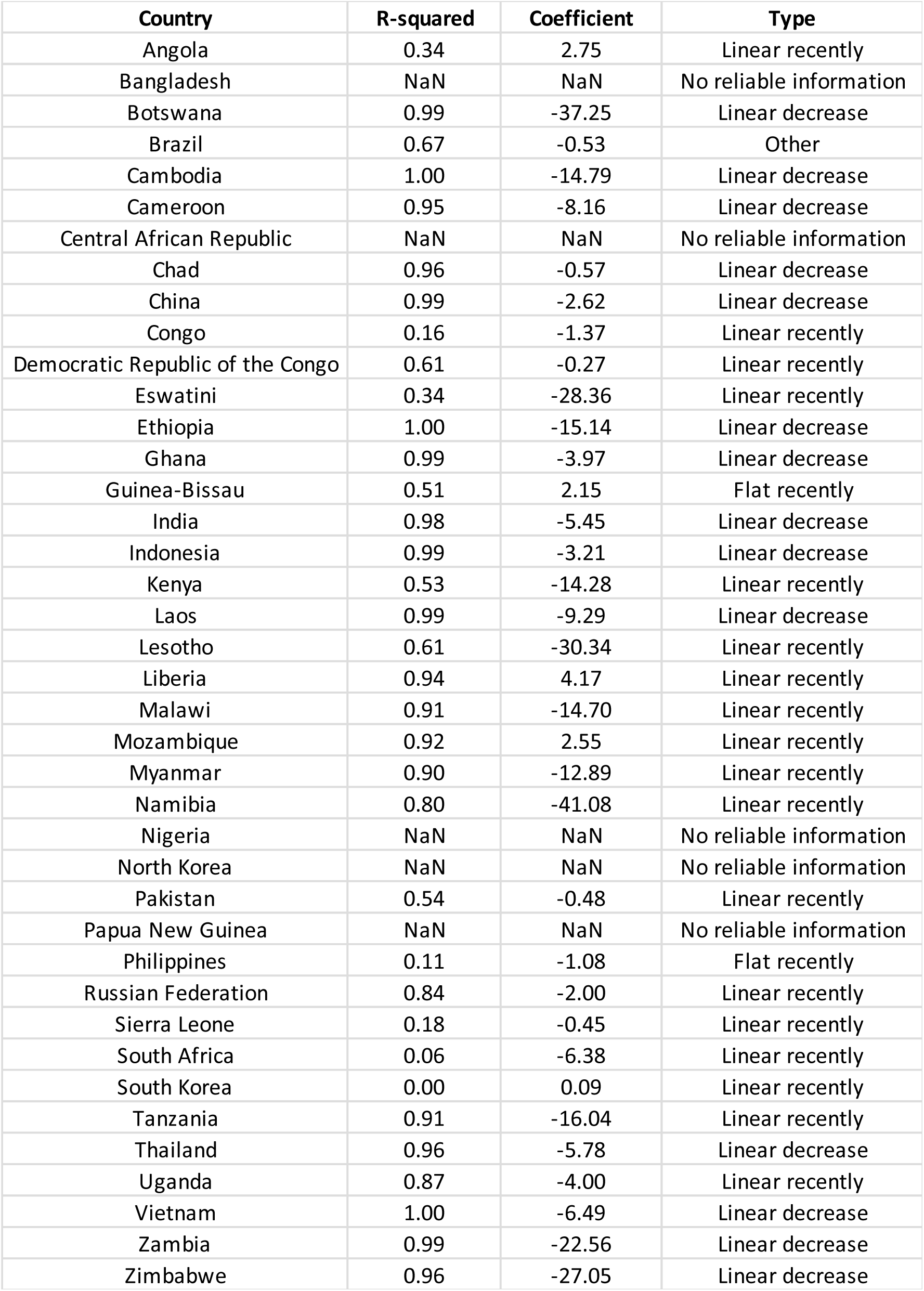
Results of linear model fit to TB incidence data for all 40 countries, 2000-2018. Table available on Figshare, DOI: 10.6084/m9.figshare.13040708.v2.

TB incidence data were obtained directly from the open-access WHO database (http://www.who.int/tb/country/data/download/en/). We have also included the data tables in the GitHub repository that hosts the code for this analysis (https://github.com/chaj1230/endTB). The variables “e_inc_100k”, “e_inc_100k_hi”, and “e_inc_100k_lo”, which represent the estimated incidence of all forms of TB per 100,000 people, its high-bound, and its low-bound, respectively, were extracted by country for the years 2000 to 2018. Population data by country were obtained from the World Bank Open Data, which includes both the reported estimates as well as World Bank projections to 2035 (https://data.worldbank.org/).

Data for country-specific HIV analyses from years 2000 to 2018 were accessed from both the WHO and the World Bank Open Data. The estimated incidence of TB/HIV coinfection per 100,000 people (“e_inc_tbhiv_100k”) was extracted from the WHO dataset. Total adult prevalence of HIV (% of population ages 15-49) and anti-retroviral (ARV) therapy coverage (% of people living with HIV) were obtained from the World Bank (https://data.worldbank.org/).

The current World Bank classifications were followed to categorise countries as low income, lower middle income, upper middle income, or high income.^14^

### Fitting linear models to the WHO data

All computational analyses were performed using R software (version 3.5.2). Scripts used to analyse and generate plots can be found on Github at https://github.com/chaj1230/endTB. The reported TB incidence from 2000 to 2018 for each country (“e_inc_100k”) was visualized using the ggplot2 package. The high and low bounds of the incidence, “e_inc_100k_hi” and “e_inc_100k_lo,” respectively, served as the confidence interval (CI). All trend lines and linear models fit to the entire time frame can be seen in Supplementary Figure 6.

We fit a linear trend to the incidence data for each of the 40 countries in our study using the lm function. We found that 5 HBCs had no change in incidence values at all, which we interpreted as the WHO treating further reports from these countries as unreliable data. We examined 21 HBCs with an R^2^ < 0.95 in more detail. Some countries displayed a linear trend in TB incidence for a more recent part of the time series; a linear model was thus fitted to this sub-section. We identified pivotal TB control efforts since 2000 and their implementation years with literature searches and considered these when selecting the years of TB incidence data to include in the linear model.

### Projection of the 2020-2035 estimates and confidence intervals

Incidence estimates for 2020-2035 were inferred for each HBC using the “predict” function in R and the model for that country. Additionally, for high HIV burden countries in Sub-Saharan Africa, the trends in TB/HIV incidence and non-HIV TB incidence were modelled separately and combined to give an overall estimate. The CI ranges for future projections were calculated by taking the average ratio of the high or low CI to the estimated incidence over the 2014 to 2018 period of the WHO data. This average ratio was then used to project the CI forward alongside the projected estimates through 2035 for each country. The minimum value for projected and target incidence was set to 10 per 100,000 people, the threshold for the WHO definition of “low incidence” of TB.^15^

The reported TB incidence values from 2000 to 2018 and the predicted trends from 2019 to 2035 using the modified linear model, along with the CI, were graphed for each country, grouped by trend.

### Comparison of the projected incidence to the End TB target incidence

For each country, the End TB target incidence was modelled by multiplying the 2015 reported incidence by 0.8 for 2020, 0.5 for 2025, 0.2 for 2030 and 0.1 for 2035. These milestones were used to precisely fit a polynomial regression of the 5^th^ degree using the glm function in R, from which target values for each year were interpolated. These targets were then compared against the predicted TB incidence values, and the net difference between the two from 2020 through 2035 was calculated. A country was concluded to miss the End TB target if the lower CI of its projected number of cases exceeded its target number of cases in 2035.

The absolute numbers of TB cases for both the projected and End TB target scenarios were calculated for the years 2020 to 2035 by multiplying the projected or target incidence per 100,000 people by the World Bank projected population size divided by 100,000.

### Analysis of HIV related trends

As untreated HIV is a known risk factor for developing TB, access to HIV antiretroviral (ARV) treatment and TB by HIV status were further analysed in a smaller cohort of 15 high HIV/AIDS burden countries in Sub-Saharan Africa. These countries included: Angola, Botswana, Cameroon, Congo, Eswatini, Kenya, Lesotho, Malawi, Namibia, Sierra Leone, South Africa, Uganda, Tanzania, Zambia, and Zimbabwe.

To examine whether the peak in the 2000s followed by a decline in TB incidence correlated to the trajectory of access to ARV therapy for HIV, the prevalence of uncontrolled HIV was calculated from ARV therapy coverage (% of people living with HIV) data and graphed against the total adult prevalence of HIV (% of population ages 15-49) for years 2000 to 2018.

TB incidence values parsed by HIV status were evaluated with “e_inc_tbhiv_100k,” “e_inc_tbhiv_100k_hi,” and “e_inc_tbhiv_100k_lo” data, which represent TB/HIV incidence and its confidence interval as estimated by the WHO. These were subtracted from their respective overall TB incidence data (“e_inc_100k,” “e_inc_100k_hi,” and “e_inc_100k_hi”) to calculate TB incidence in HIV uninfected people. Confidence intervals, projections, calculations of the numbers of projected cases, and comparisons against End TB goals were conducted as explained with the overall TB incidence data, resulting in three separate analyses: overall TB, TB in HIV-infected people (TB/HIV), and TB in HIV-uninfected people. Countries that had been projected to be on track to meet targets for overall TB but failed to meet targets in either of these two sub-analyses by HIV status were concluded to miss End TB targets. As TB incidence in HIV uninfected people was not directly reported by WHO, and it was calculated by subtracting the incidence in HIV infected people from total incidence, there were no confidence intervals available for this estimate or its projection.

## Results & Discussion

### Trends in TB incidence in high burden countries

The reported TB incidence of 14 of the 40 analysed HBCs was well-described by a linear model (R^2^ > 0.95) with a negative slope (Table 1). This reflects a remarkably steady decline in TB incidence in these countries: Botswana, Cambodia, Cameroon, Chad, China, Ethiopia, Ghana, India, Indonesia, Laos, Thailand, Vietnam, Zambia, and Zimbabwe (Figure 1). While the consistency of decline was similar among these countries, the rates of decrease (in incidence per 100,000) varied between *−*0.57 ∗ *year* in Chad and *−*37.25 ∗ *year* in Zimbabwe (full results in Table 1). This wide range is likely driven by the varying epidemiology of TB in these countries (HIV associated or not), as well as the overall stage of TB control, with faster absolute decreases being achieved in countries with higher incidence.

**Figure 1.**
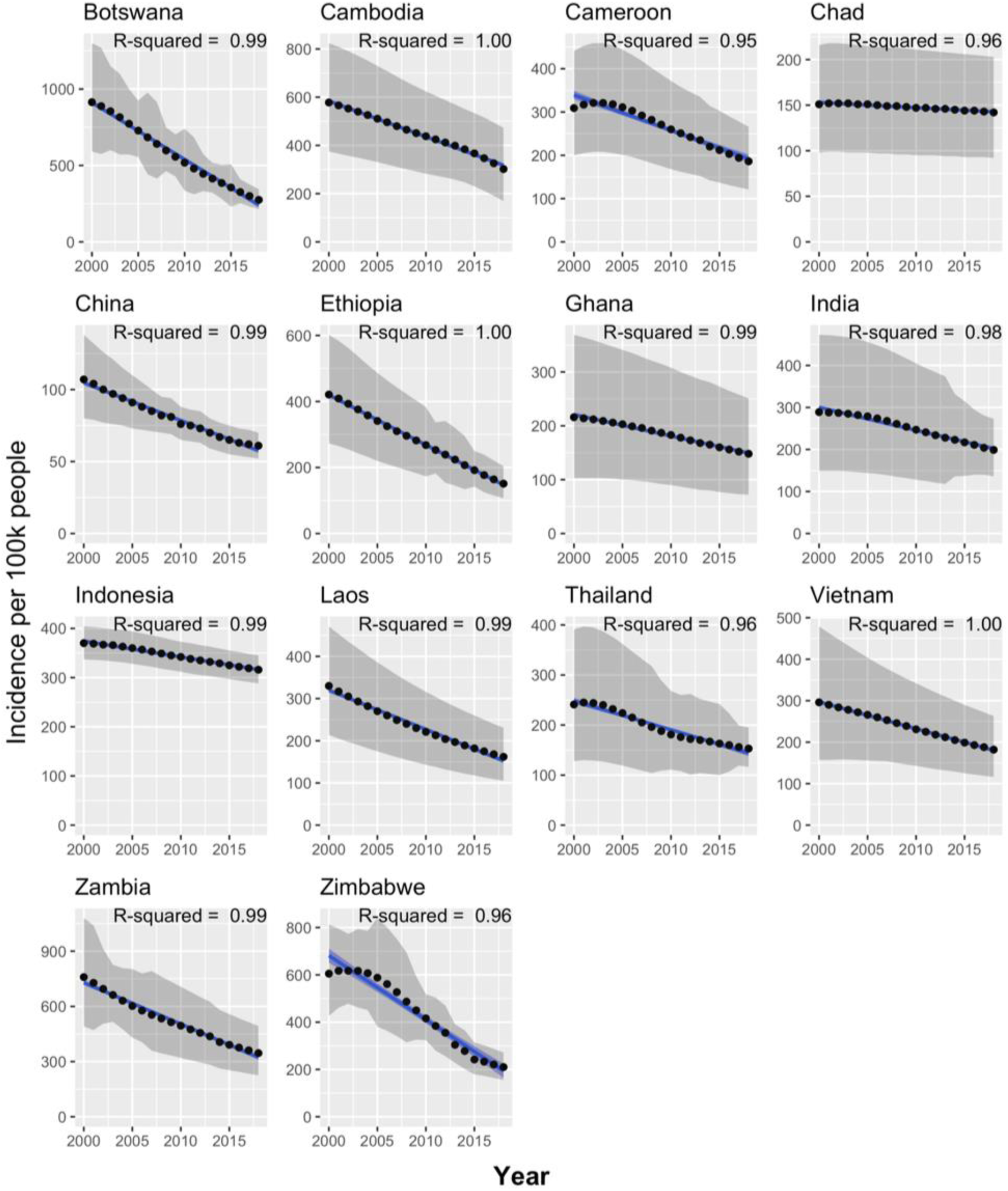
TB incidence in countries with a linear trend, 2000-2018. Black points represent WHO estimated incidence, grey shaded areas represent the WHO high and low bounds for that estimate, blue lines display the fit of the linear model, and the blue shaded areas represent the confidence interval of the linear model.

There were 12 HBCs characterised by a distinctive TB incidence trend of a peak during the 2000s (Figure 2). All countries are Sub-Saharan African with a high prevalence of HIV: Angola, Cameroon, Congo, Eswatini, Kenya, Lesotho, Malawi, Namibia, Sierra Leone, South Africa, Tanzania and Zimbabwe. The HIV prevalence in these countries ranges from 1.5% in Namibia to 23.6% in Kenya (average = 11.7%). As uncontrolled HIV/AIDS is a well-established risk factor for TB, the HIV treatment coverage in these countries for the period 2000 to 2018 was examined (Supplementary Figure 3). This revealed that 7 of these countries (Angola, Cameroon, Eswatini, Lesotho, Namibia, Sierra Leone, and South Africa) had also experienced a peak in the prevalence of uncontrolled HIV in the 2000s, while 5 of the countries (Congo, Kenya, Uganda, Tanzania, and Zimbabwe) have had steady declines in uncontrolled HIV since 2000. The incidence of TB/HIV coinfection revealed that declines in TB incidence in 8 countries (Eswatini, Kenya, Lesotho, Malawi, Namibia, South Africa, Tanzania, and Zimbabwe) were primarily driven by corresponding declines in the incidence of TB in people infected with HIV, while HIV-uninfected TB incidence decreased relatively slowly (Figure 5 & 6).

**Figure 2.**
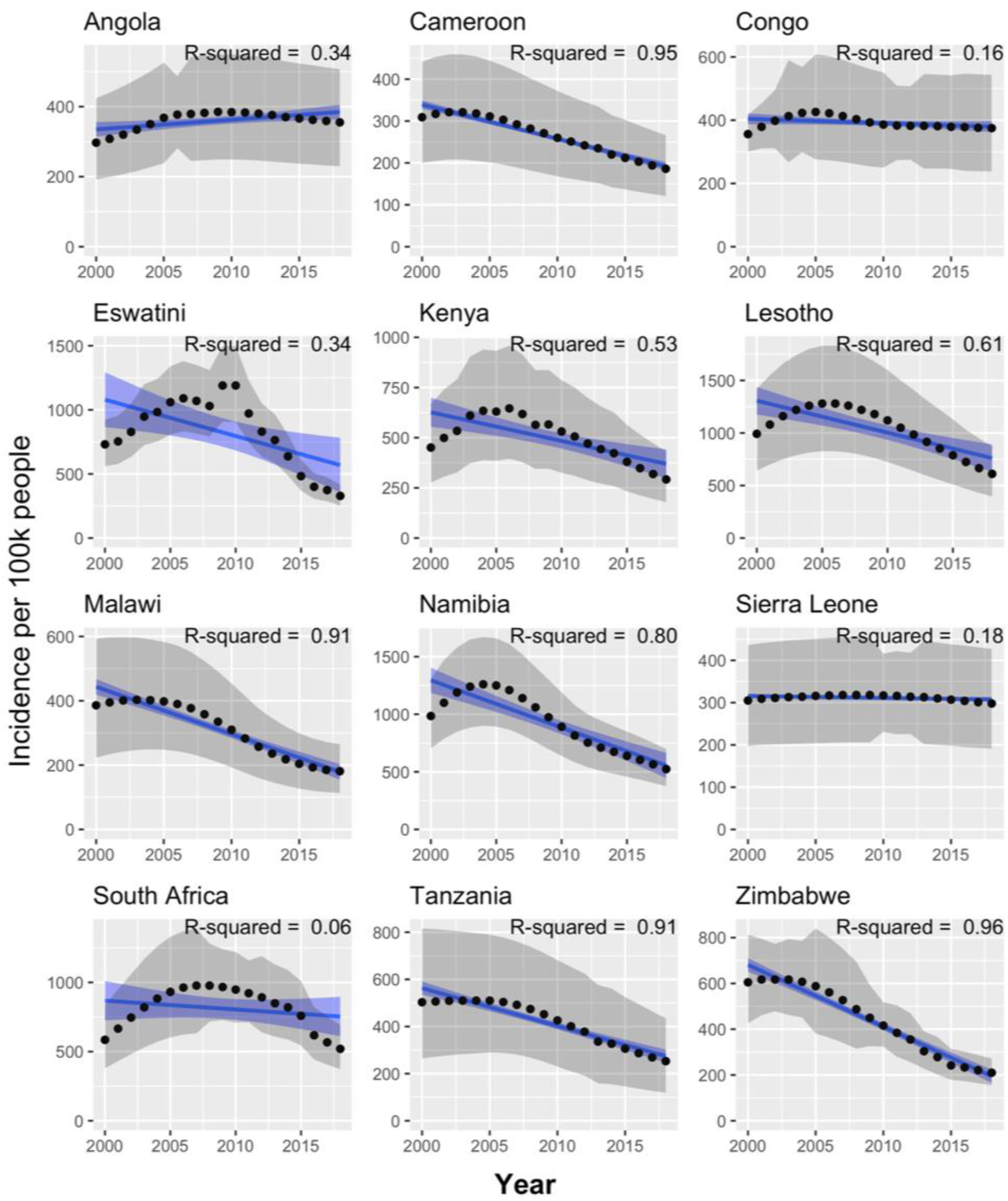
TB incidence in countries whose TB incidence peaked in the 2000s and then declined, 2000-2018. Figure colours as per Figure 1.

**Figure 3.**
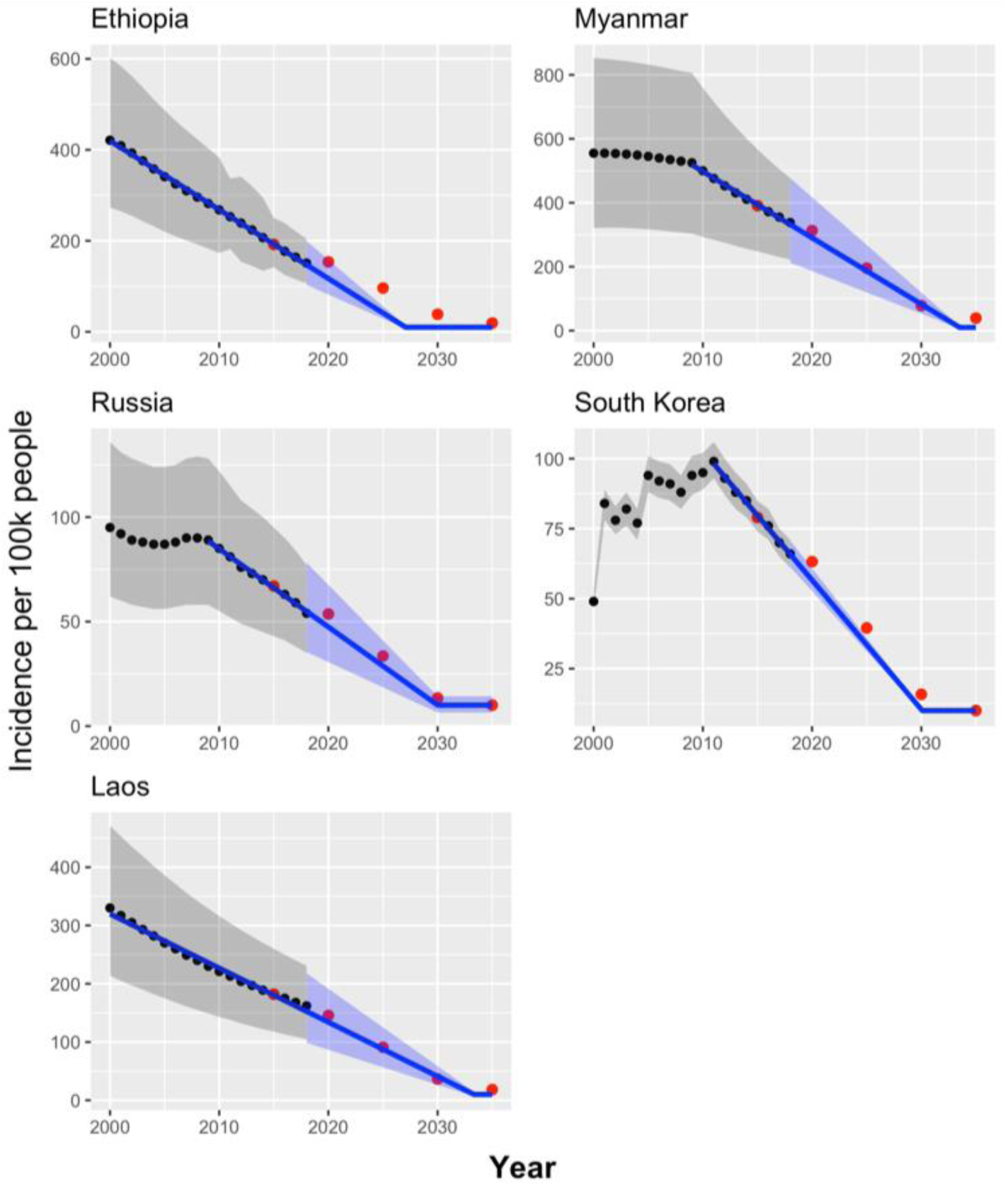
TB incidence (2000-2018) and projections from this study (2019-2035) in countries without high HIV burden that are predicted to meet End TB targets. Figure colours as per Figure 1. The red points represent the WHO End TB benchmark targets for incidence.

**Figure 4.**
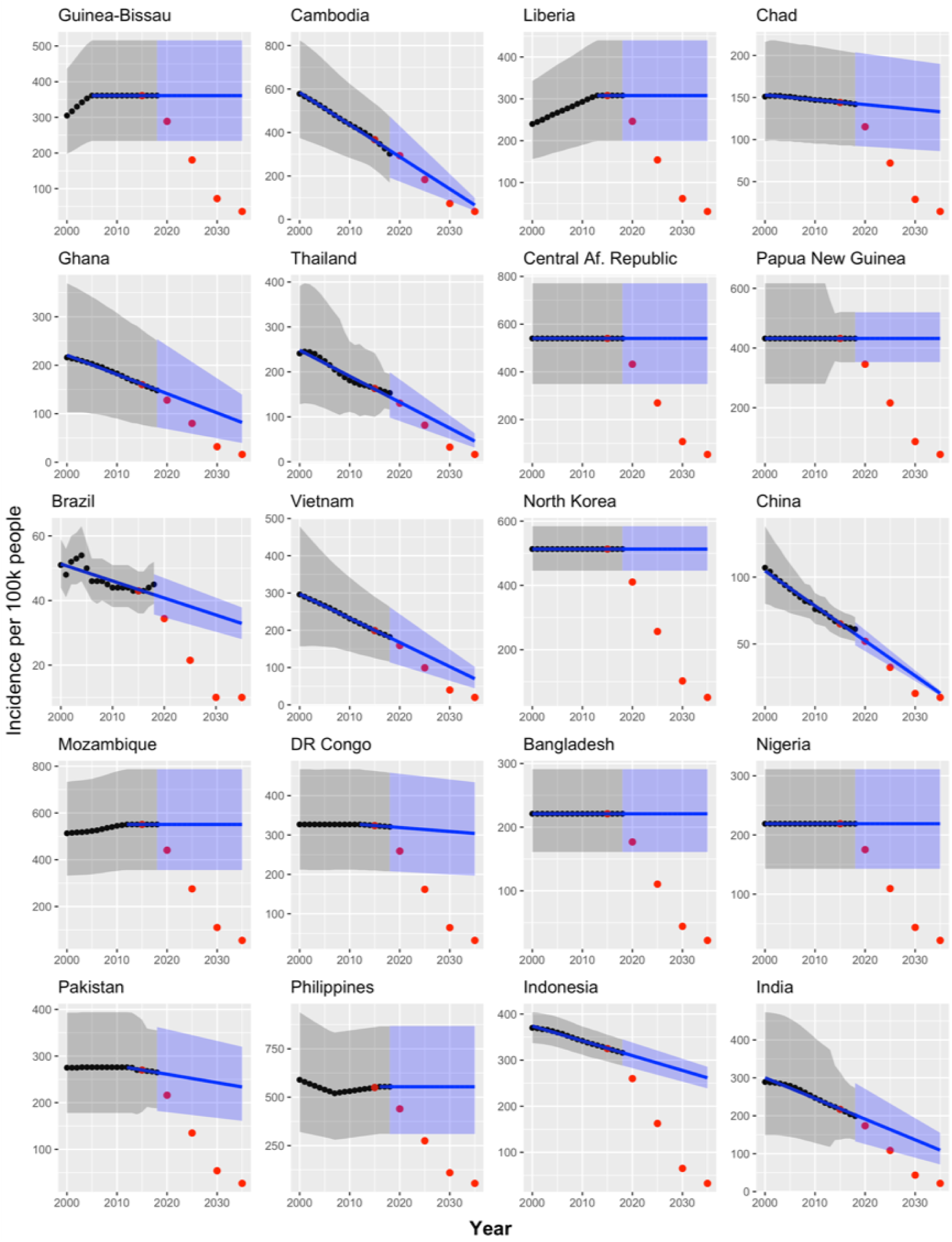
TB incidence (2000-2018) and projections from this study (2019-2035) in countries without high HIV burden that are predicted to miss End TB targets. Figure colours as per Figure 1. The red points represent the WHO End TB benchmark targets for incidence.

**Figure 5.**
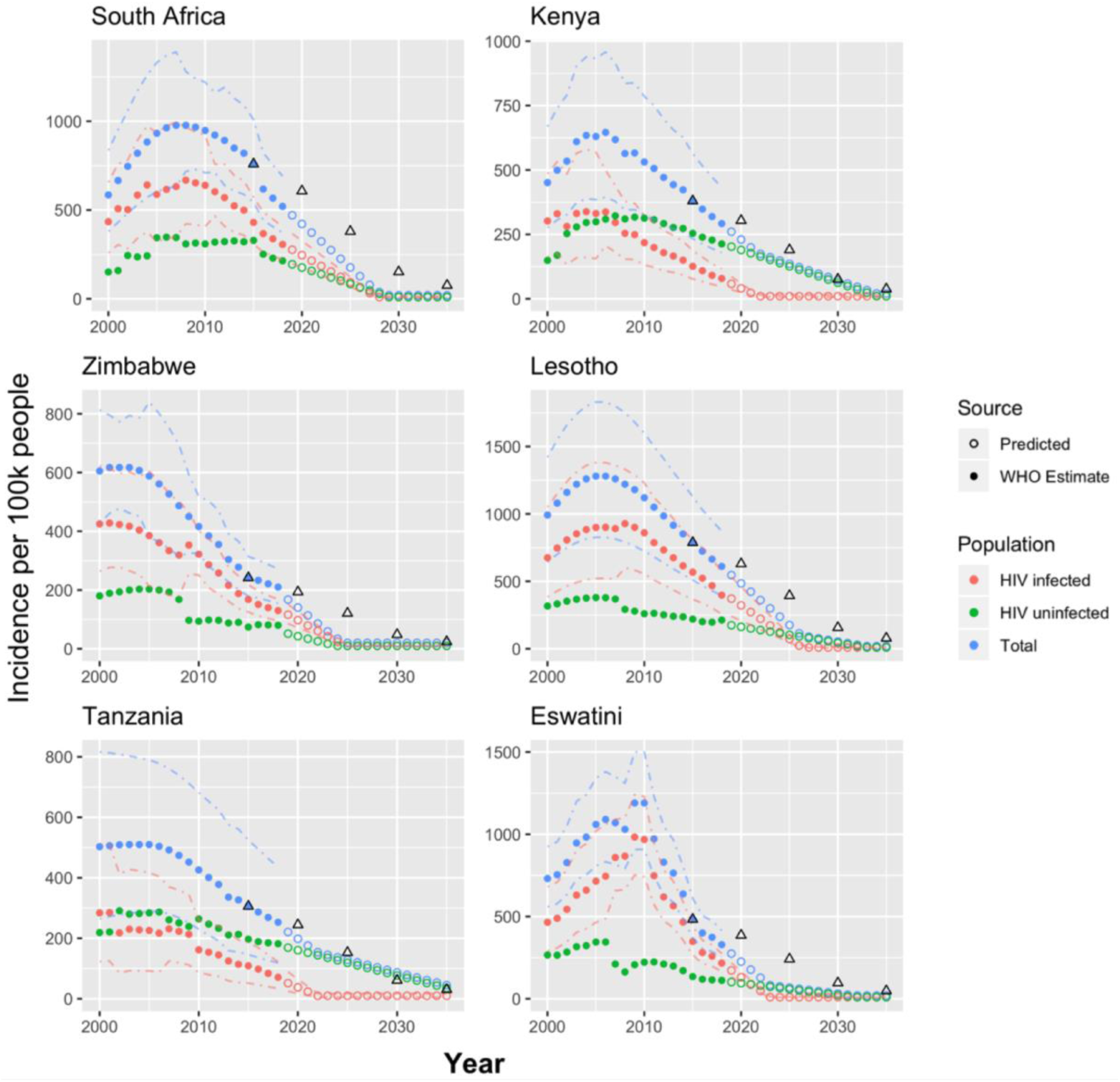
TB incidence by HIV infection status (2000-2018) and predictions from this study (2019-2035) in high HIV burden countries predicted to meet End TB targets. Dot-dash lines represent confidence intervals on the WHO estimates, which were only provided for total TB and TB in HIV infected people. Dashed lines represent confidence intervals on projections, which were only available for the HIV infected population. Colour of dashed and dot-dash lines denotes Population. Black triangles represent the End TB targets.

Despite the global decrease in TB incidence,^1^ 4 HBCs (Angola, Guinea-Bissau, Liberia and Mozambique), had overall trends of increasing TB incidence between 2000 and 2018 (Table 1 & Supplementary Figure 1). There was a trend of decreasing incidence in Angola from 2009, but not enough to compensate for the rapid increase between 2000 and 2009. For Guinea-Bissau, Liberia, and Mozambique, the increases were approximately linear until 2005, 2013, and 2012 respectively, after which the numbers became fixed. This suggests that data quality deteriorated at this time, leading the WHO to assume constant TB incidence onwards.^16^ While a definite cause cannot be determined for this unexpected trend of increasing incidence, one possibility is that Angola, Liberia, and Mozambique have experienced civil wars until 2002, 2003, and 1992, respectively, with ongoing conflict pervasive in these regions. The poor data quality from 3 of these 4 countries, recently too low to even be included, and the wide CIs associated with previous estimates indicate that interpretations of incidence from these countries warrant caution.

Bangladesh, the Central African Republic, Nigeria, North Korea, and Papua New Guinea reported the same TB incidence from 2000 to 2018 (Supplementary Figure 2). One explanation for this, suggested by the 2009 WHO report on TB surveillance, is an assumption that incidence is constant in the absence of sufficient data collected by a country.^16^ Thus, the lack of change in estimated TB incidence indicates the lack of effective and/or consistent reporting in these 5 HBCs from 2000 to 2018. For instance, Bangladesh has had considerable changes in case finding efforts leading to fluctuations in notification rates, and thus its TB incidence has been assumed to be flat until more data becomes available.^16^ The lack of sufficient surveillance to infer incidence in these countries is worrying, considering that they have a combined population of nearly 400 million people and include two of the most densely populated mega-cities in the world, Dhaka and Lagos.

An examination of the 8 HBCs for which the trends have not already been discussed (Supplementary Figure 4) reveals that an individualized approach, with consideration of the health systems, national policies for TB control, war and conflict, HIV prevalence, and reporting methods, among other factors, is necessary to situate and better understand the trends in these countries. As there are broad CIs associated with estimates from some of these countries, the figure was also generated without CIs to enable clearer viewing of the estimated incidence (Supplementary Figure 5). The Philippines exhibited a singular V-shaped curve of TB incidence with a linear decrease until the year 2008, followed by a slower, linear increase, then three years of no reported change. The Philippines dramatically improved its TB surveillance system in 2008, which may account for this reported increase in TB cases,^17^ but the subsequent lack of change from 2016 to 2018 indicates that the WHO deemed reporting too unreliable to modify the estimated incidence. This further highlights the disconnect between the reported and actual TB incidence in lower income countries without adequate reporting systems. The Democratic Republic of the Congo (DRC), Myanmar, and Pakistan all had stable or unchanged incidence until 2013, 2010, and 2013 respectively, followed by either dramatic (Myanmar) or modest (DRC and Pakistan) declines. Myanmar was internationally isolated until 2011 following a landmark general election in 2010,^18^ and the subsequent changes in TB incidence may be due to changes in the reporting or management of cases in the country. The DRC is characterised by chronic political instability and is one of the least developed countries in the world, which is reflected in its modest decline in TB incidence between 2013 and 2018. Reported numbers from Pakistan likely suffer from inadequate surveillance and may be far from capturing the true incidence of TB; its number of notified TB cases increased significantly from 34,066 in 2001 to 248,115 in 2008, ^19^ yet no evident change is indicated in the WHO TB incidence data. Russia experienced growing TB incidence between 2005 and 2008, likely reflecting increased problems with MDR-TB in the country.^20^ Uganda has seen its rate of decline in TB incidence slow between 2000 and 2018, partly due to increasing incidence of TB in HIV uninfected people (Supplementary Figure 3 & Figure 6).

**Figure 6.**
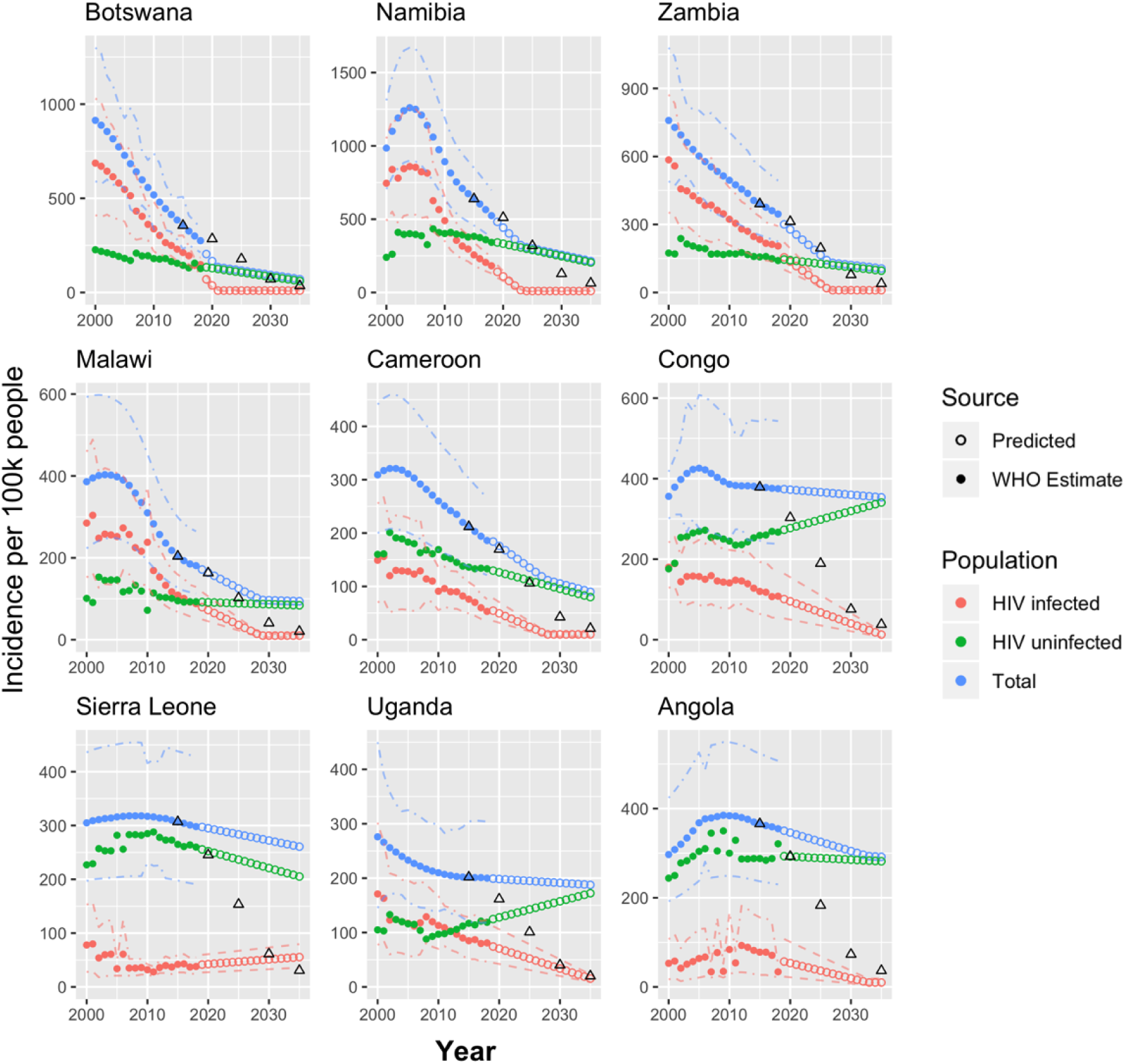
TB incidence by HIV infection status (2000-2018) and predictions from this study (2019-2035) in high HIV burden countries predicted to miss the End TB targets. Figure colours and annotations as per Figure 5.

The trend of TB incidence in South Korea was particularly noteworthy. Although South Korea is not on the WHO high burden list globally, it has the highest incidence of TB among the 36 OECD (Organisation for Economic Co-operation and Development) countries.^21^ Korea’s remarkable economic growth in recent decades and robust national TB control efforts have been noted as instrumental to its management of TB from a prevalence (per 100,000 people) of 5,168 in 1965 to 767 in 1995 to 77 in 2016.^21,22^ The sustained decrease in TB incidence from 2011 onwards reflects the almost immediate effectiveness of new strategies implemented by the Korean government to combat TB. These include the 2011 implementation of the “New 2020 Plan for Early TB Elimination” with targeted governmental efforts on early detection, heightened intensive care efforts, and a dramatic three-fold increase in the budget for TB control. Many initiatives have followed throughout the decade, including the Five Year Plan for TB Elimination (2013-2017), the TB-free Korea programme (2016), and intensified screenings and expansion of insurance to cover all medical expenses for TB, even the living costs for families of MDR-TB patients.^21,22^ South Korea has had unprecedented economic growth in a short time span, government-mandated universal health care, and substantial efforts by the state to control TB.^21-23^ In this regard, South Korea has the potential to be adopted as a model for the trajectories of high TB burden countries currently undergoing rapid economic growth.

Twenty of the 40 countries displayed partial, rather than continuous, linear decreases in incidence starting from a year subsequent to 2000. We projected the recent linear trends of these countries, defined empirically on a case by case basis for each country (Table 1).

### Which countries are on track to meet the WHO End TB targets?

Next, we compared the TB incidence trends in these 40 HBCs to the decreases necessary to meet the WHO End TB target of a 90% reduction in incidence between 2015 and 2035. Because of the dramatic impact of HIV on the epidemiology of TB, we analysed 15 high HIV burden countries from Sub-Saharan Africa separately. For these 15 countries, TB incidence rates in HIV infected and uninfected people were modelled individually to account for different trends in the two populations.

Of the 25 countries without a high burden of HIV, only 5 are on track to meet the End TB targets (Table 2, Figure 3): Ethiopia, Laos, Myanmar, Russia, and South Korea. These countries are projected to have a combined 972,000 fewer cases than the End TB targets from 2020 to 2035, with individual countries ranging from 4,303 (Laos) to 760,545 (Ethiopia) cases. As for the reasons underlying this success, in the case of Ethiopia, it is notable that Dr. Tedros Adhanom was a much-lauded Minister of Health from 2005 to 2012, with impressive performance across a range of health indicators.^24^ Laos also has had a strong record of TB control. It met the TB related Millennium Development Goal and the Stop TB target of halving the prevalence of TB between 1990 and 2015, partly driven by the availabilities of Directly Observed Therapy Short-course (DOTS) treatments in 95% of its health centres and anti-TB medication at no cost.^25^ The steps South Korea has taken to control its TB incidence have been detailed above and similarly entail interventions that increase both the quantity and quality of TB treatment.

**Table 2.**
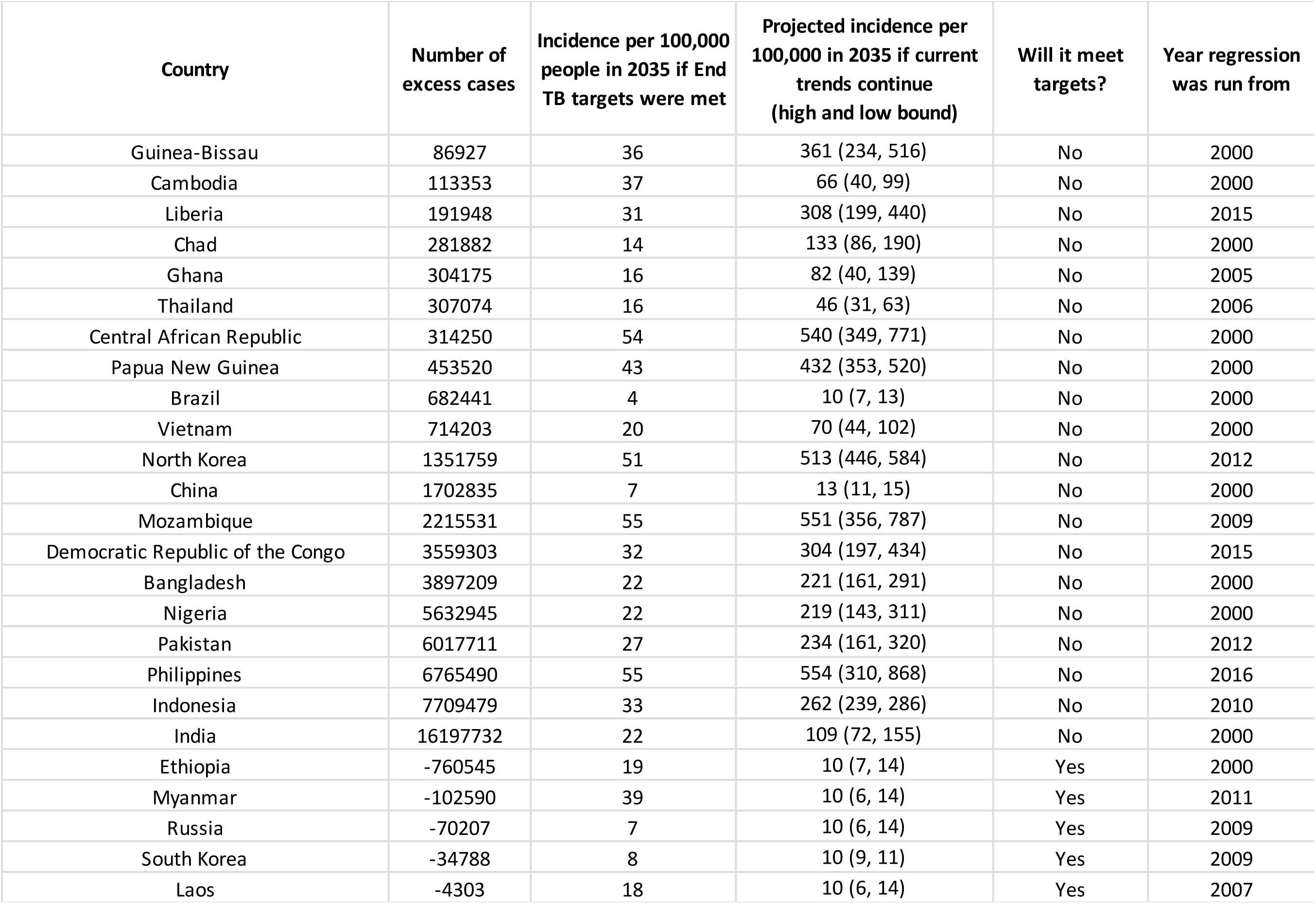
Assessment of End TB targets in 25 countries without high HIV burden. Table available on Figshare, DOI: 10.6084/m9.figshare.13040708.v2.

The remaining 20 low HIV burden countries are projected to miss the targets (Table 2, Figure 4) with a combined excess of 58.5 million cases of TB. More than half of these excess cases are accounted for by only 3 countries: India, Indonesia, and the Philippines. Progress toward reducing TB incidence in these countries has been hampered by decentralised health systems, public-private mixed models of health service leading to patients lost along the diagnosis-treatment pathway, and poor surveillance reflected in large CIs or unchanging incidence values in the WHO statistics. Four countries (Brazil, Cambodia, China, and Thailand) will come close to meeting the targets, but the End TB goal in 2035 is not within the lower bound of their projected incidence (Table 2). Despite narrowly missing the targets, Brazil is projected to achieve effective elimination by 2035 (incidence of 10 cases per 100,000 people), and China will have only 13 cases per 100,000 people, which would still represent a major global health achievement. Progress toward effective elimination of TB in Brazil may have been bolstered by the provision of universal free TB care and a range of other social programmes to improve health.^26^ China has made steady improvements in a variety of health indicators, and its DOTS coverage reached 100% in 2005.^27^

Of the 15 countries with high HIV burdens, 6 are on track to meet the End TB targets (Table 3, Figure 5): Eswatini, Kenya, Lesotho, South Africa, Tanzania, and Zimbabwe. These 6 countries will go beyond the reductions required to meet the End TB targets by a combined total of 2 million cases averted, varying between 22,596 (Eswatini) and 1.4 million (South Africa) cases. The large number of cases averted in Sub-Saharan Africa, and particularly in South Africa, is consistent with an existing evaluation of End TB milestone for 2020.^13^ The downwards step change in South Africa’s TB incidence in 2016 suggests a change in reporting methodology at this time that contributed towards the number of cases averted against the 2015 benchmark. All of these countries, except Tanzania, are on track to achieve effective elimination of TB in both HIV infected and uninfected people by 2035. Ensuring continuation of these great gains will require focused socio-political and economic commitments. A particularly striking example of success is Eswatini, which declared a national TB emergency in 2011, tripled the number of diagnostic sites, increased TB management units, and instituted active case finding.^28^ The effectiveness of this response is clearly reflected in Eswatini’s trajectory.

**Table 3.** Assessment of End TB targets in 15 countries with high HIV burden, separated by HIV infection. Table available on Figshare, DOI: 10.6084/m9.figshare.13040708.v2.

| Country | HIV infected |  | HIV uninfected |  | Overall |  | Will it meet targets? | Number of excess cases |
| --- | --- | --- | --- | --- | --- | --- | --- | --- |
|  | TB Incidence per 100,000 people in 2035 if End TB targets were met | Projected TB incidence per 100,000 people in 2035 (high and low bound) | TB Incidence per 100,000 people in 2035 if End TB targets were met | Projected TB incidence per 100,000 people in 2035 (high and low bound) | TB Incidence per 100,000 people in 2035 if End TB targets were met | Projected TB incidence per 100,000 people in 2035 (high and low bound) |  |  |
| South Africa | 43 | Effective elimination | 16 | Effective elimination | 39 | Effective elimination | Yes | -1424402 |
| Kenya | 13 | Effective elimination | 12 | Effective elimination | 20 | Effective elimination | Yes | -348484 |
| Zimbabwe | 17 | Effective elimination | Effective elimination | Effective elimination | 24 | Effective elimination | Yes | -131941 |
| Lesotho | 57 | Effective elimination | 20 | Effective elimination | 31 | Effective elimination | Yes | -51109 |
| Tanzania | 11 | Effective elimination | 38 | 35 (17, 61) | 64 | 45 (27, 71) | Yes | -23109 |
| Eswatini | 35 | Effective elimination | 33 | Effective elimination | 76 | Effective elimination | Yes | -22596 |
| Botswana | 21 | Effective elimination | 14 | 61 (45, 79) | 36 | 71 (55, 89) | No* | -10290 |
| Namibia | 26 | Effective elimination | 25 | 205 (147, 272) | 38 | 215 (157, 282) | No* | 25976 |
| Zambia | 23 | Effective elimination | 27 | 96 (62, 137) | 31 | 106 (72, 147) | No* | 56628 |
| Malawi | 11 | Effective elimination | Effective elimination | 84 (52, 123) | 48 | 94 (62, 133) | No | 157789 |
| Cameroon | Effective elimination | Effective elimination | 14 | 80 (52, 114) | 21 | 90 (62, 124) | No | 235378 |
| Congo | 12 | 13 (10, 21) | 26 | 341 (236, 463) | 38 | 354 (246, 484) | No | 242721 |
| Sierra Leone | Effective elimination | 56 (36, 80) | 22 | 205 (132, 293) | 79 | 261 (168, 373) | No | 245897 |
| Uganda | Effective elimination | 15 (10, 23) | 10 | 172 (103, 259) | 20 | 188 (113, 283) | No | 1088780 |
| Angola | Effective elimination | Effective elimination | 29 | 282 (201, 368) | 37 | 292 (211, 378) | No | 1249365 |

There are 9 high HIV burden countries currently projected to miss the End TB targets (Table 3, Figure 6). In an initial analysis of the overall trend (i.e. without separating HIV infected and HIV uninfected), 3 of these countries (Botswana, Namibia, and Zambia) had been projected to be on track to meet the End TB targets.This speaks to the slower control of TB in HIV uninfected individuals in these countries compared to great progress in the TB/HIV syndemic. Of particular note is Botswana, where 10,290 *fewer* cases of TB are projected compared to End TB targets overall. This counterintuitive result is likely because Botswana is projected to dramatically exceed the End TB targets until TB in HIV infected people is effectively eliminated in 2021, after which progress toward targets will slow dramatically. Sierra Leone is the only country to display a trend of increasing incidence of TB in HIV infected people, while the Congo and Uganda both show decreases in overall TB and TB/HIV but an increase in the incidence of TB in HIV uninfected people. Greater national and international focus on the TB programmes in these countries is required to ensure that these alarming trends are reversed, especially as they have thus far been hidden within the values of overall incidence.

Of the 30 HBCs on the WHO list, the 2019 WHO TB report had estimated that 11 are on track to meet the 2020 milestone of a 20% reduction in incidence: Cambodia, Ethiopia, Kenya, Lesotho, Myanmar, Namibia, Russia, South Africa, Tanzania, Zambia, and Zimbabwe.^1^ Of these 11, our projections show 8 are on track to continue and meet the targets through 2035, with Namibia and Zambia both projected to miss the targets when HIV infected and uninfected TB incidences are modelled separately. Cambodia, the third discrepancy, is projected to come close to the End TB targets with only 113,000 excess cases between 2020 and 2035.

One weakness of this study is the underlying assumption of a linear trend to TB incidence. On balance, we believe this is a reasonable assumption given that 14 countries had an R^2^ > 0.95 when their incidence data from 2000 to 2018 were fit to a linear model. However, it still remains a simplification, especially for countries for which only a subset of the time series was used to fit the linear model. In addition, while we took projected changes in population sizes into account when calculating the absolute number of projected cases, changes in risk factors such as smoking, diabetes mellitus, malnutrition, poverty, poor air quality, and age profiles of the population were not used to directly model the projected TB incidence. Nevertheless, HIV, the most significant risk factor and comorbidity, was evaluated. Another limitation is that as MDR-TB has different epidemiological trends to non-MDR-TB, countries on the WHO list for MDR-TB,^1^ but not overall TB, were excluded. Similar analyses may be of interest for these 10 countries (Azerbaijan, Belarus, Kazakhstan, Kyrgyzstan, Peru, Moldova, Somalia, Tajikistan, Ukraine, and Uzbekistan) to determine their progress towards the WHO End TB targets by 2035. As noted above, there are numerous countries with low quality data indicating that the current incidence estimates do not reflect the reality of TB incidence in the population. These countries have still been included in our analyses, as to exclude them may falsely suggest that there is no problem with TB in those countries. While they detract from the accuracy of our projected estimates, they do not change the overall message, either per country or globally.

### The trajectory of TB in the wake of COVID-19

While novel and emerging viruses have the potential to wreak death and disruption across the globe, the number of COVID-19 deaths as of writing has yet to reach the average number of lives lost to TB every year. The global incidence of TB has been decreasing since 2000, with an average decline of 1.6% per year from 2000 to 2018.^1^ As revealed by our analysis, Sub-Saharan African countries, many with HIV as a major driver of TB, have achieved some of the largest percentage reductions in TB incidence. However, these gains in TB control are threatened by the spread of SARS-CoV-2 in the ongoing pandemic.

COVID-19 poses both direct and indirect consequences for TB control. Recent literature indicates an association between pre-existing TB with increased susceptibility to and severity of COVID-19.^29-31^ One clinical study suggests that comorbidity with TB manifests atypical radiological features in individuals diagnosed with COVID-19.^32^ Although a clear causal relationship between the two is debated,^33^ the similarities in their initial physiological symptoms and shared risk factors (including poverty, overcrowding, and air pollution) mean that *M. tuberculosis* and SARS-CoV-2 are two intertwined infectious agents whose coinfection likely leads to worse prognosis than either one alone.^29-31,33^

Indirectly, the ongoing pandemic redirects resources and attention away from TB, as well as from HIV and malaria. Restrictions in international commerce that limit access to necessary medications, shifts in manufacturing of TB diagnostic tests to COVID-19 tests, and reallocation of financial and human resources towards COVID-19 are only a few of many concerns for the future of TB control. A June 2020 survey by the Global Fund, which provides 69% of all international financing for TB, found that 78% of TB and 85% of HIV programmes face disruptions in care delivery due to COVID-19.^35^ The similarity of initial symptoms between COVID-19 and TB further exacerbates the situation, fuelling stigmatisation and reluctance of health workers to deliver comprehensive treatment.

Modelling studies carried out since COVID-19 emerged predict a regression of global TB control, including significant increases in TB mortality, ranging from 13% in 2020 to 20% through 2025.^36,37^ One study from the Stop TB Partnership and USAID (United States Agency for International Development) predicts 6.3 million excess TB cases globally between 2020 and 2025 from COVID-19, assuming a 3-month lockdown and a 10-month recovery, as an *under*estimate.^38^ With an end to the pandemic uncertain, decades-long gains in TB control may be reversed as a repercussion of the pandemic unless strong corrective actions are taken.

## Conclusions

Our projections reveal that 29 of the 40 analysed countries will not meet the WHO End TB targets if current trends continue, with an excess of 61,792,010 cases between 2020 and 2035 compared to if the targets were met. Seven of the 11 countries projected to meet targets are in Sub-Saharan Africa. Three high HIV burden countries that had appeared on track to meeting targets when only overall incidence was projected were revealed to miss targets when TB incidence in HIV uninfected people were modelled separately. This reveals that as the number of people with uncontrolled HIV diminishes, the rate of decrease in TB incidence is likely to slow. The ongoing COVID-19 pandemic is also expected to disrupt the projected trajectories of TB incidence. Therefore, our prediction that 11 of the 40 countries analysed will meet the End TB targets may still be a significant overestimate, and more directed programmes for TB control should be concentrated in these regions with consideration for country-specific contexts and successful measures in peer countries.

## Data Availability

All data were obtained from the open-access WHO database and the World Bank. Scripts used to analyse and generate plots can be found on Github. Tables can be assessed on Figshare.

http://www.who.int/tb/country/data/download/en/

https://data.worldbank.org/

https://github.com/chaj1230/endTB

https://doi.org/10.6084/m9.figshare.13040708.v2

## Author contributions

JC carried out data gathering, analyses, interpretation, and writing. GET contributed to interpretation and writing. PMA conceived the study, carried out data gathering, analyses, interpretation, and writing.

## Conflict of interest, Funding Statement & Ethics

The authors declare no conflict of interest. The funder had no role in the design or interpretation of the study and had no role in the decision to publish. All data were from secondary sources in the public domain, and ethics approval was thereby not required.

## Acknowledgements

We would like to acknowledge Hannah Clapham for helpful discussions and Duy Nguyen Manh for assistance with R code.

## Supplementary Figures

**Figure S1.**
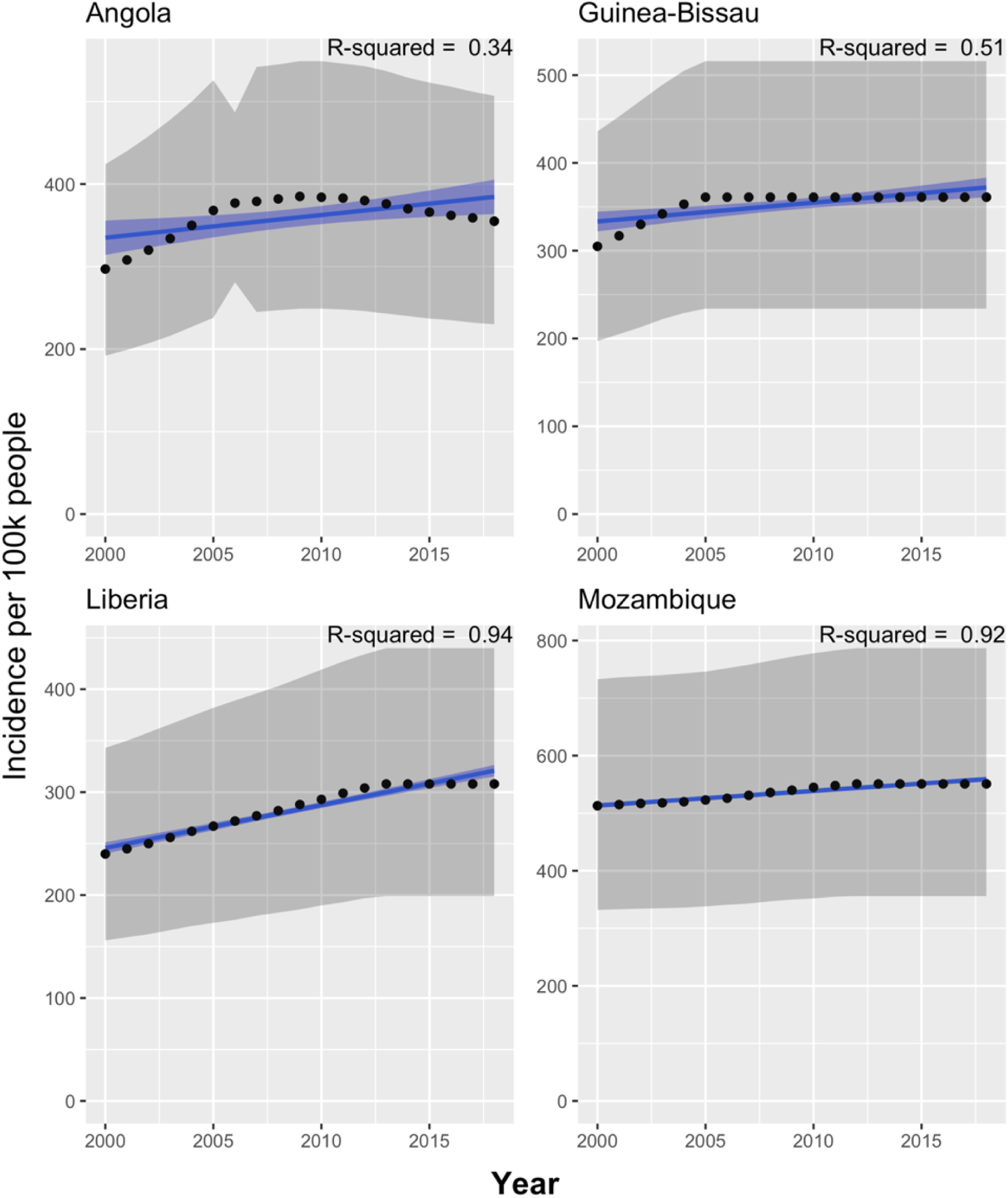
Countries with increasing TB incidence, 2000-2018. TB incidence over time for countries with a trend line showing overall increase. Figure colours as per Figure 1.

**Figure S2.**
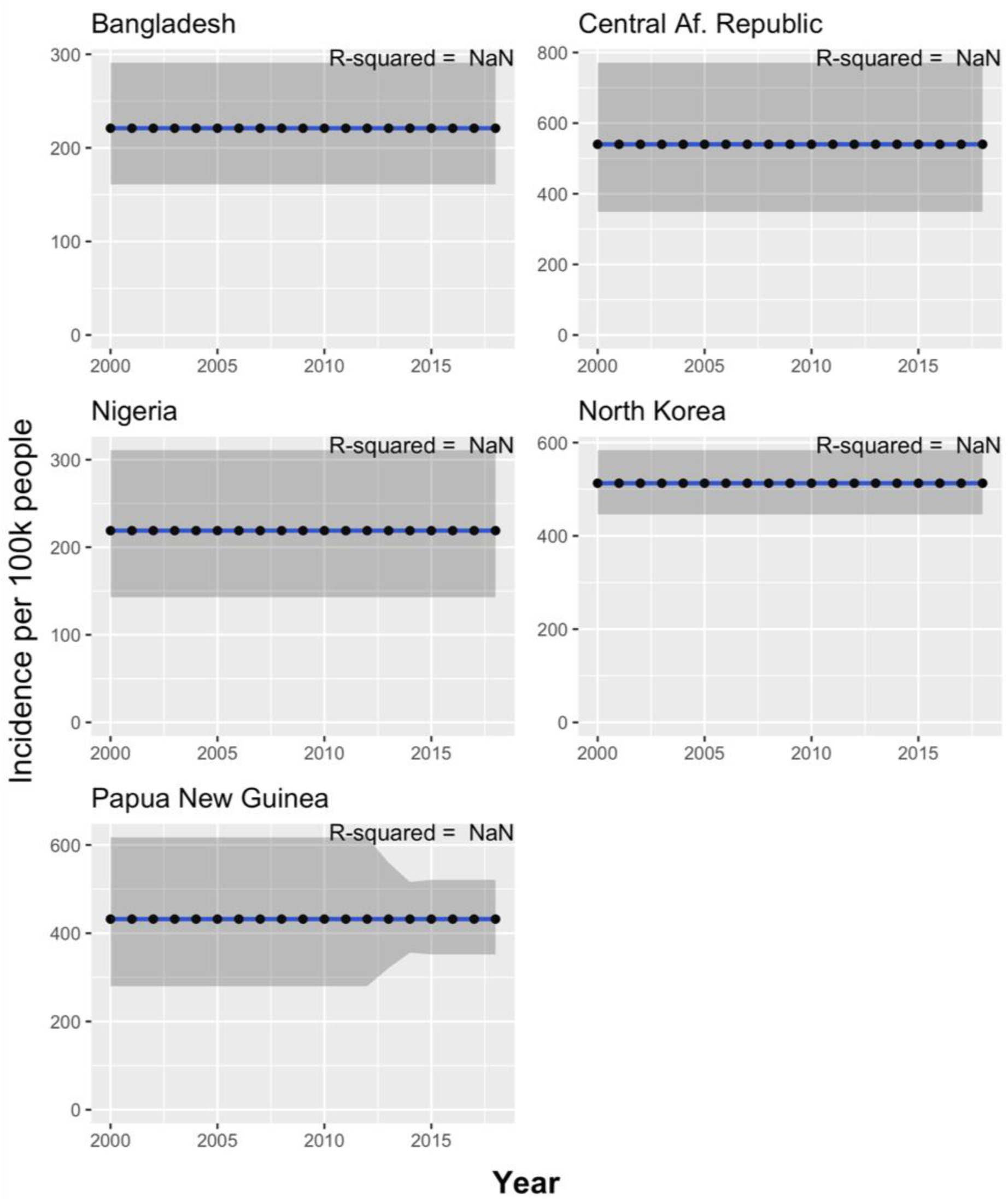
Countries with unchanging TB incidence, 2000-2018. TB incidence over time for countries with no change in their TB incidence. Figure colours as per Figure 1.

**Figure S3.**
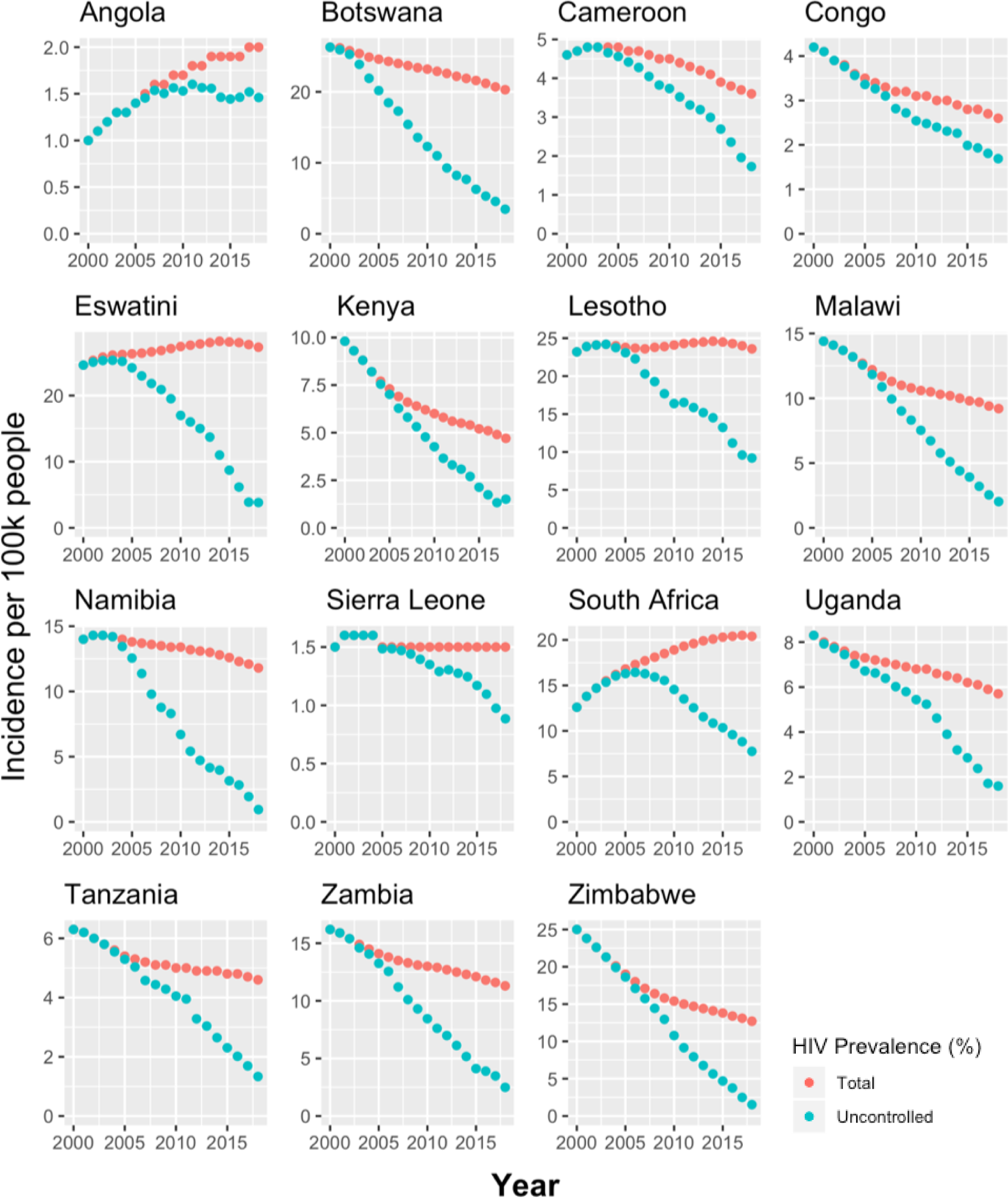
Total and uncontrolled HIV prevalence of high TB/HIV burden countries, 2000-2018.

**Figure S4.**
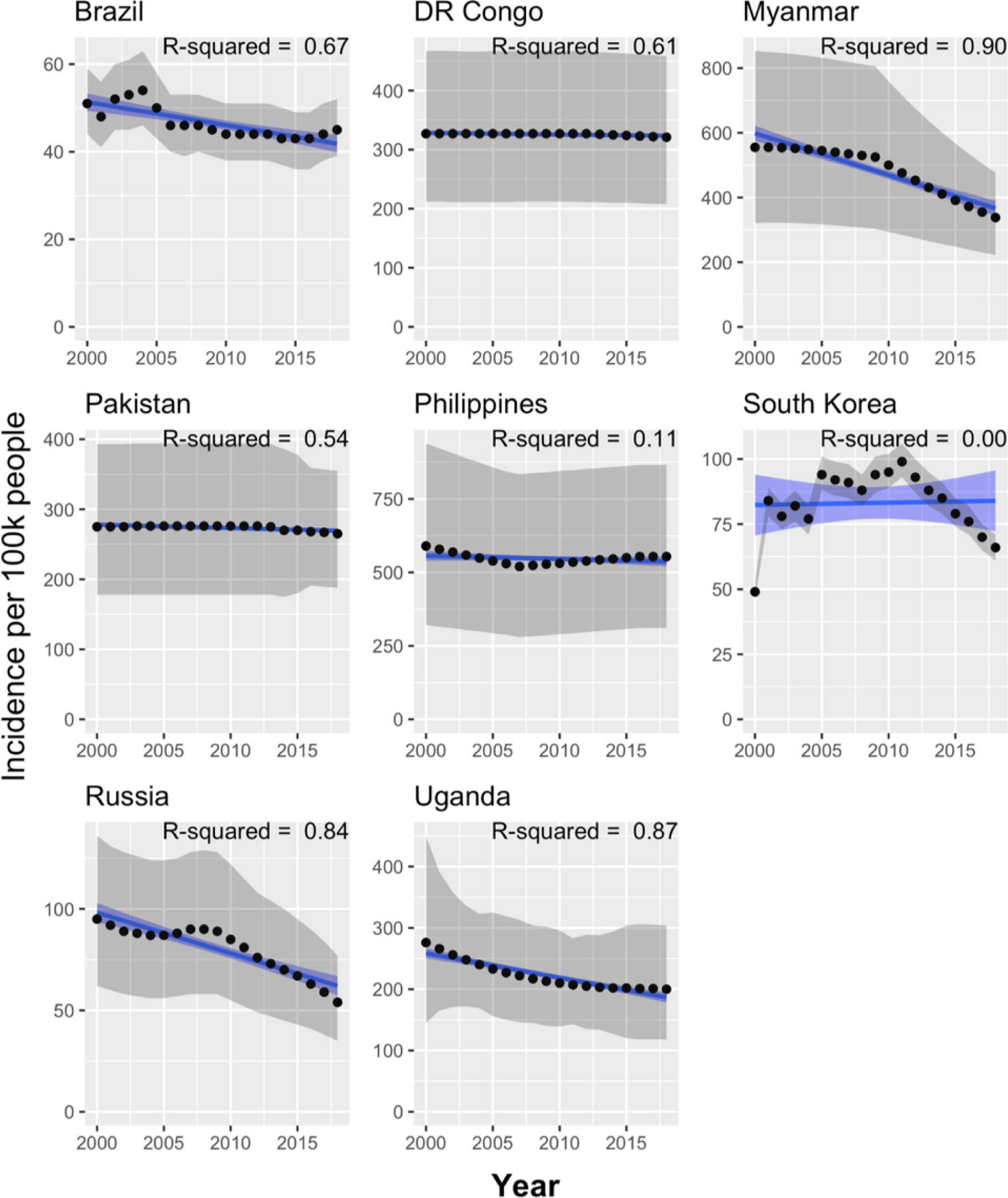
Countries with “other” TB incidence, 2000-2018. TB incidence over time for countries that did not group with other countries’ trajectories. Figure colours as per Figure 1.

**Figure S5.**
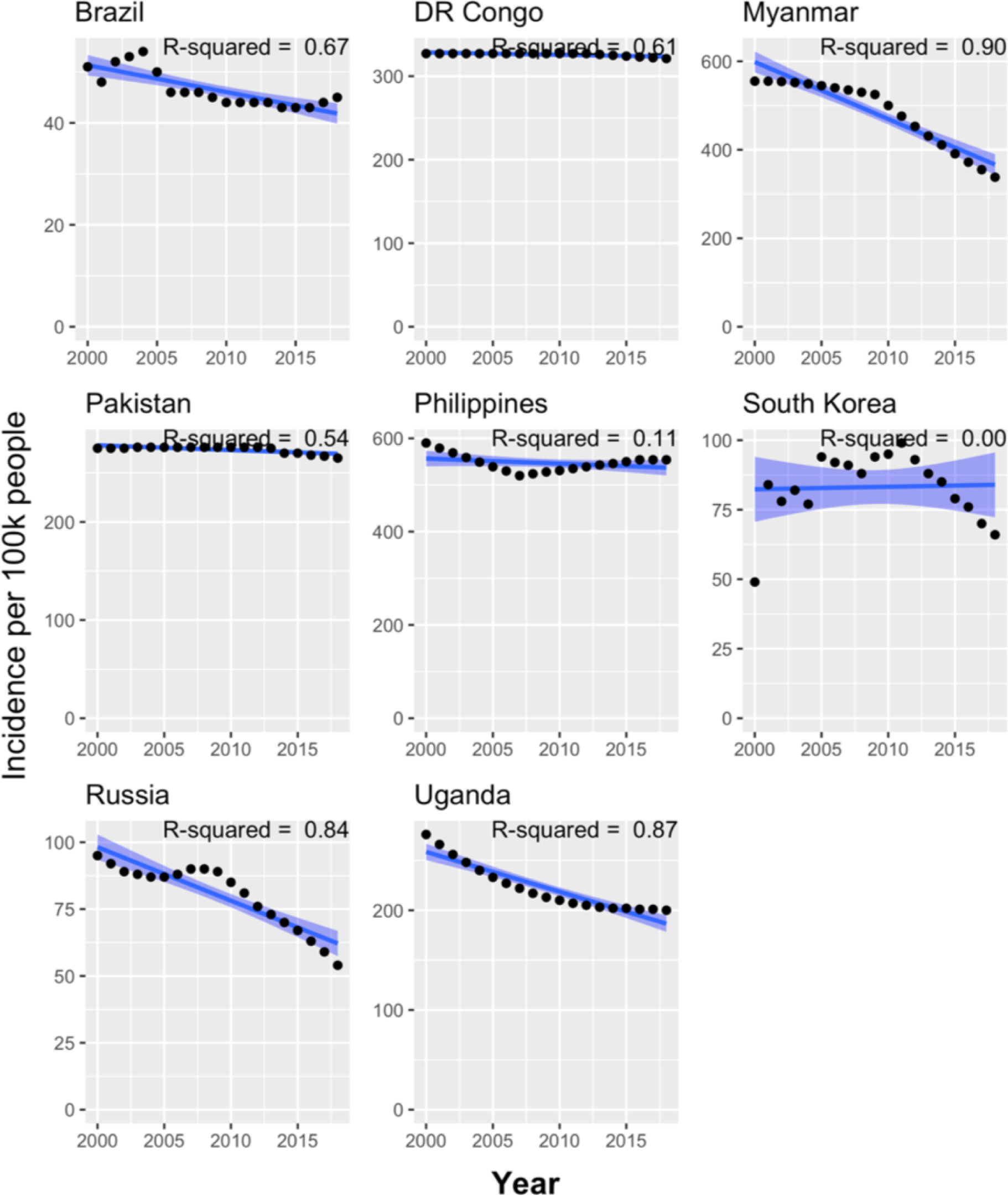
Countries with “other” TB incidence, no confidence intervals, 2000-2018. TB incidence over time for countries that do not group neatly with other countries’ trajectories, plotted without confidence intervals. Figure colours as per Figure 1.

**Figure S6.**
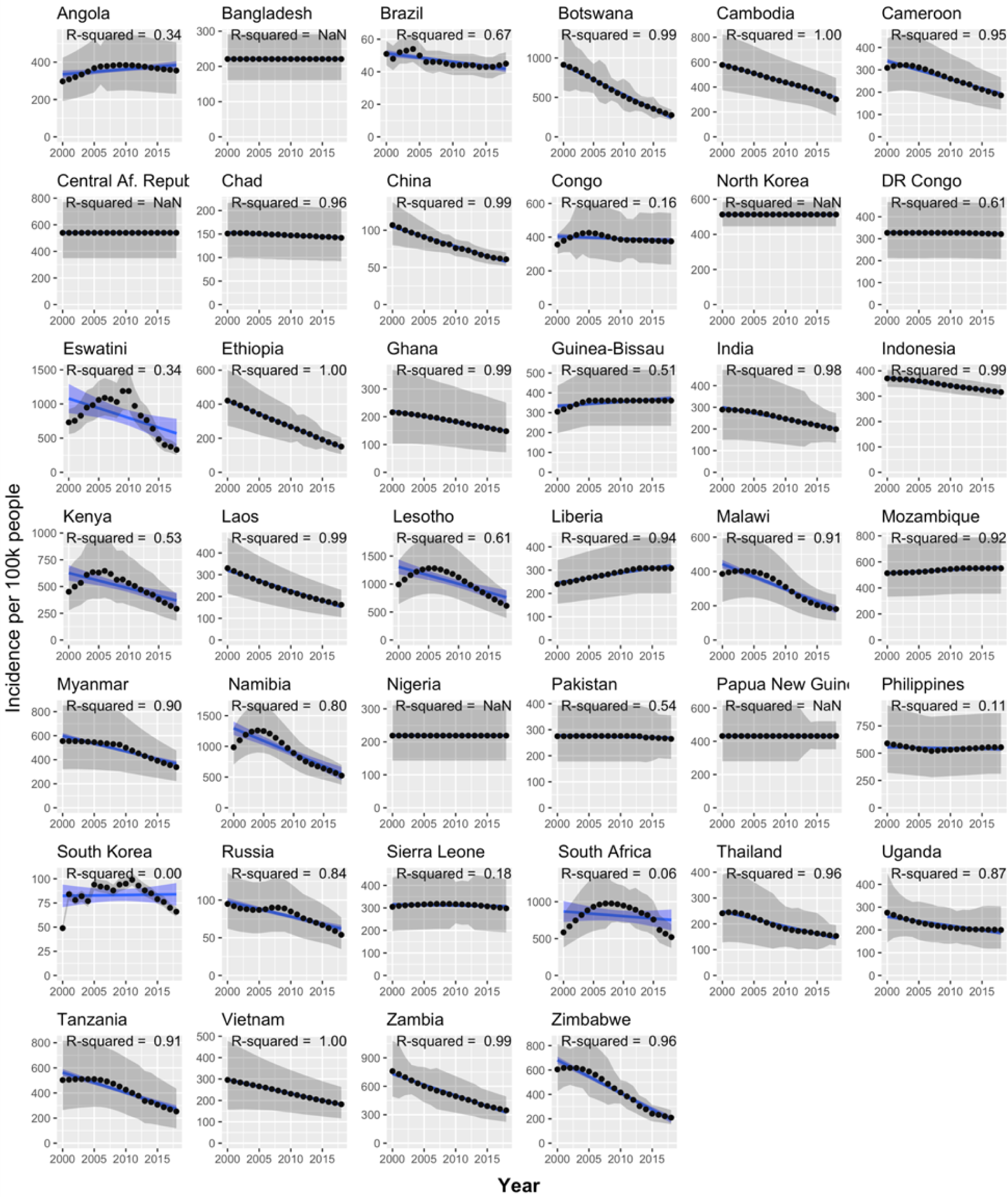
TB incidence in all 40 countries, 2000-2018. Figure colours as per Figure 1.

